# School Neighborhood Deprivation is Associated with a Higher Prevalence of Hypertension

**DOI:** 10.1101/2024.04.09.24305590

**Authors:** Elizabeth A. Onugha, Ankona Banerjee, Kenneth J. Nobleza, Duc T. Nguyen, Omar Rosales, Abiodun Oluyomi, Jayna M. Dave, Joshua Samuels

## Abstract

**Background:** Hypertension, a major modifiable risk factor for cardiovascular disease, is increasing among adolescents. While socioeconomic factors have been shown to contribute to hypertension risk in adults, similar data in children is lacking. We examined the association between socioeconomic characteristics of school neighborhoods and the prevalence of hypertension in adolescents.

**Methods:** School neighborhood socioeconomic status (SES) was assessed via the area deprivation index (ADI), a composite measure of area-level socioeconomic deprivation. Using data from a school BP surveillance program implemented in the Houston metropolitan area, we performed a cross-sectional analysis using both univariate and multivariable regression analyses.

**Results:** Among 21,392 students aged 10 to 19 years during the 2000 through 2017 academic years, the prevalence of sustained hypertension was 2.6%. Although 85% of students attended schools located in neighborhoods in quartiles 1 (Q1) and 2 (Q2) (higher ADI represents more deprivation), Hispanics and African Americans were overrepresented in the schools located inside more deprived neighborhoods. The highest sustained hypertension rate (4.6%) was observed among students attending Q 3 and Q4 schools compared to 2.3% in Q 1 and Q2 schools (p < 0.001). Multivariable regression analysis showed that being male, obese, and attending school in a disadvantaged neighborhood were significantly associated with an increased prevalence of hypertension.

**Conclusions:** Attending school in neighborhoods with lower SES may account for the increased hypertension prevalence in minority adolescents. Characteristics of school neighborhoods play an important role in the disparities observed in hypertension prevalence, providing direct opportunities to encourage improved dietary intake and physical activity, both of which are important determinants of hypertension.

## INTRODUCTION

Primary hypertension is the leading cause of hypertension in adolescents with an estimated prevalence of 3-5% in the United States.^1–3^ This condition exhibits a trajectory into adulthood, ^4^ children can have neurocognitive deficits and develop evidence of target organ damage specifically left ventricular hypertrophy and arterial stiffening, highlighting the significance of hypertension. ^5–8^ The development of primary hypertension involves a complex interplay of genetic, environmental, and behavioral factors.^2^ Several risk factors for primary hypertension have been identified, including race, ethnicity, prematurity, low birth weight, obesity, stress, increased sodium intake, and physical inactivity.^9–16^ Moreover, there is a growing recognition of the impact of socioeconomic deprivation on hypertension risk and blood pressure (BP) control in adults.^17–19^

Numerous studies have demonstrated that social determinants at the neighborhood level contribute to disparities in disease prevalence and outcomes, perpetuating health inequities.^20–23^ Living in low-income neighborhoods is associated with an increased prevalence of hypertension and poor BP control among adults, ^17^ due to adverse neighborhood characteristics such as high crime rates, lack of access to healthy food options, poor walkability, and limited healthcare access. ^24–27^ While this association is less clear in children, parental education and insurance status have been associated with hypertension risk in this population.^20,25,26^ Additionally, research suggests that characteristics of school neighborhoods influence the risk of obesity and diabetes among students. ^27, 28^

A study using Delaware Medicaid data found an association between neighborhood poverty level and hypertension diagnosis in youth.^18^ However, there has been no investigation into the relationship between a validated measure of school neighborhood deprivation and hypertension prevalence in the U.S. The close link between public education and housing due to residential zoning regulations makes the school neighborhoods a good surrogate for a student’s residential environment. Nearly 40 percent of funding for public education in the U.S. comes from local property taxes, leading to significant socioeconomic disparities between school districts.^29^ In the current analysis, we investigated the association between school-level neighborhoods and hypertension risk among adolescents in the Houston metropolitan area of Texas in the US. We hypothesized that there would be a higher prevalence of hypertension in schools located in disadvantaged neighborhoods compared to schools in affluent neighborhoods.

## METHODS

### Study Setting and Population

We used the Houston school-based BP screening data collected by the Houston Pediatric and Adolescent Hypertension Program at the University of Texas McGovern Medical School. These data were collected between 2000 and 2017 from schools that were selected via convenience sampling to mirror the racial/ethnic distribution of the Houston metro population. Over the study period, 21,392 students from 28 participating schools were enrolled. The study procedure has been reported elsewhere.^3,30^ All enrolled students were eligible to participate, and screening procedures occurred during physical education classes. The school BP screening study was approved by the Institutional Review Board at the University of Texas Health Science Center at Houston, as well as local school district institutional Review Boards where required. Individual consents were obtained when applicable from the legal guardian following local school district policy. From the school BP dataset, we obtained individual-level demographic data collected using questionnaires at the time of BP measurement including age, sex, and race/ethnicity. Additionally, BP, weight and height information were retrieved from measurements conducted by trained study personnel.^3, 30^ BMI percentiles depicting conventional pediatric categories (<85th percentile: normal, 85th–94th percentile: overweight, ≥95th percentile: obese) were based on the 2000 Centers for Disease Control and Prevention growth charts.

### Outcome

Our primary outcome was the prevalence of sustained hypertension. Using the Houston Pediatric and Adolescent Hypertension Program protocol, all first BP measurements were discarded, and a mean of the remaining readings was calculated to categorize each student’s BP status as:

1. Normal BP: mean systolic BP (SBP) and diastolic BP (DBP) <120/80 AND <90^th^ percentile for age, height, and sex at first screening,
2. Elevated BP: Mean SBP or DBP ≥ 90th percentile, but ≤ 95^th^ percentile (or 120- 129/80 mm Hg for subjects ≥13 years), or
3. Hypertension: Mean SBP or DBP ≥95th percentile (or ≥130/80 mm Hg for subjects’ ≥13 years) according to the 2017 American Academy of Pediatrics Clinical Practice Guidelines.^31^

Students with abnormal BP measurements on the initial screening day had repeated BP measurements on up to two additional occasions to confirm the presence of sustained hypertension per AAP clinical practice guidelines. Subsequent BP measurements were performed within two months of the initial reading to reduce potential confounders from changes in body habitus.

### Area Deprivation Index

The Area Deprivation Index (ADI) is a composite measure of neighborhood socioeconomic deprivation that relies on 17 indicators from the U.S. Census database. These indicators include educational attainment, employment rates, average household income, and homeownership rates.^32, 33^ Each school was geocoded based on its physical address to determine the neighborhood (i.e. U.S. census tract) that contains the school. For analysis, we first computed the ADI scores for all the census tracts in Texas. Then, the scores were reclassified into quartiles; where quartile 1 (Q1) represented the least deprived neighborhoods while quartile 4 (Q4) represented the most deprived neighborhoods. We implemented a weighted ADI calculation to accurately reflect the socioeconomic status of the school’s catchment area. This involved creating a 0.5-mile radius buffer around each school and assessing what percentage of this buffer was occupied by individual census tracts. The weighting of the ADI for each school was then adjusted based on the proportional representation of these census tracts within the buffer. This method ensures that the ADI value assigned to a school incorporated the ADI scores of all the tracts that intersect the 0.5-mile buffer.

### Statistical Analyses

Descriptive data were reported as frequencies and proportions for categorical variables and as median and interquartile range (IQR) for continuous variables. Differences in student characteristics between ADI quartile groups were determined by the chi-square or Fisher’s exact tests for categorical variables and the Kruskal-Wallis test for continuous variables. Bar charts were used to depict the distributions of hypertension by ADI quartiles. Logistic regression modeling was used to determine factors associated with hypertension (Yes vs. No) and depicted by forest plots. In our logistic regression analyses, clustering variance estimators were applied for schools. The selection of variables for the multivariable models was conducted using the least absolute shrinkage and selection operator (LASSO) method with the cross-validation (CV) selection option.^34^ Briefly, all variables used in the univariable analysis were assessed by the Lasso program, which suggested good models that included the variables with the highest probability of being a risk factor. The likelihood ratio test further reduced model subsets. During the modeling process, the potential risk factors were discussed with clinicians to ensure the biological plausibility of the selected covariates. To avoid over fitting, some variables which were significant in the univariate analysis, but insignificant in multivariable modeling were not selected in the final model if their exclusion did not affect the diagnostic performance of the final model. All analyses were conducted using Stata version 18.0 (StataCorp LLC, College Station, TX, USA).^35^ A p-value of <0.05 was considered statistically significant.

## Results

### Study Population

Characteristics of the study population have been previously described.^3,30^ Briefly, of the 21,392 students included in this analysis, 48.6% were male and median age was 13 years. The sample comprised primarily of White (33.2%), African American (24.8%), and Hispanic (33.7%) children (Table 1). Regarding quartile distribution, 10,013 students (46.8%) attended schools in Q1, 8,356 students (39.1%) in Q2, 800 students (3.7%) in Q3 and 2,223 students (10.4%) in Q4 (Figure 1A). African American and Hispanic students were over-represented in Q3 and Q4. The median weighted ADI for quartiles were 12.3 (Q1), 35.7 (Q2), 68.1 (Q3) and 94.4 (Q4) (Figure S1). The overall median weighted ADI of the schools sampled was 25.9 (Table 1). Among the sampled students, 15,505 (72.5%) had normal BP, 2,822 (13.2%) had elevated BP, 3,065 (14.3%) had hypertension, as defined by the 2017 American Academy of Pediatric Clinical Practice Guidelines. There was no difference in the age of students who had normal BP compared to those with hypertension; median age was 13 years. There was a male predominance in the hypertension group (57.3%), consistent with previous findings (Table 2). The prevalence of sustained hypertension in our cohort was 2.6%.

**Figure 1.**
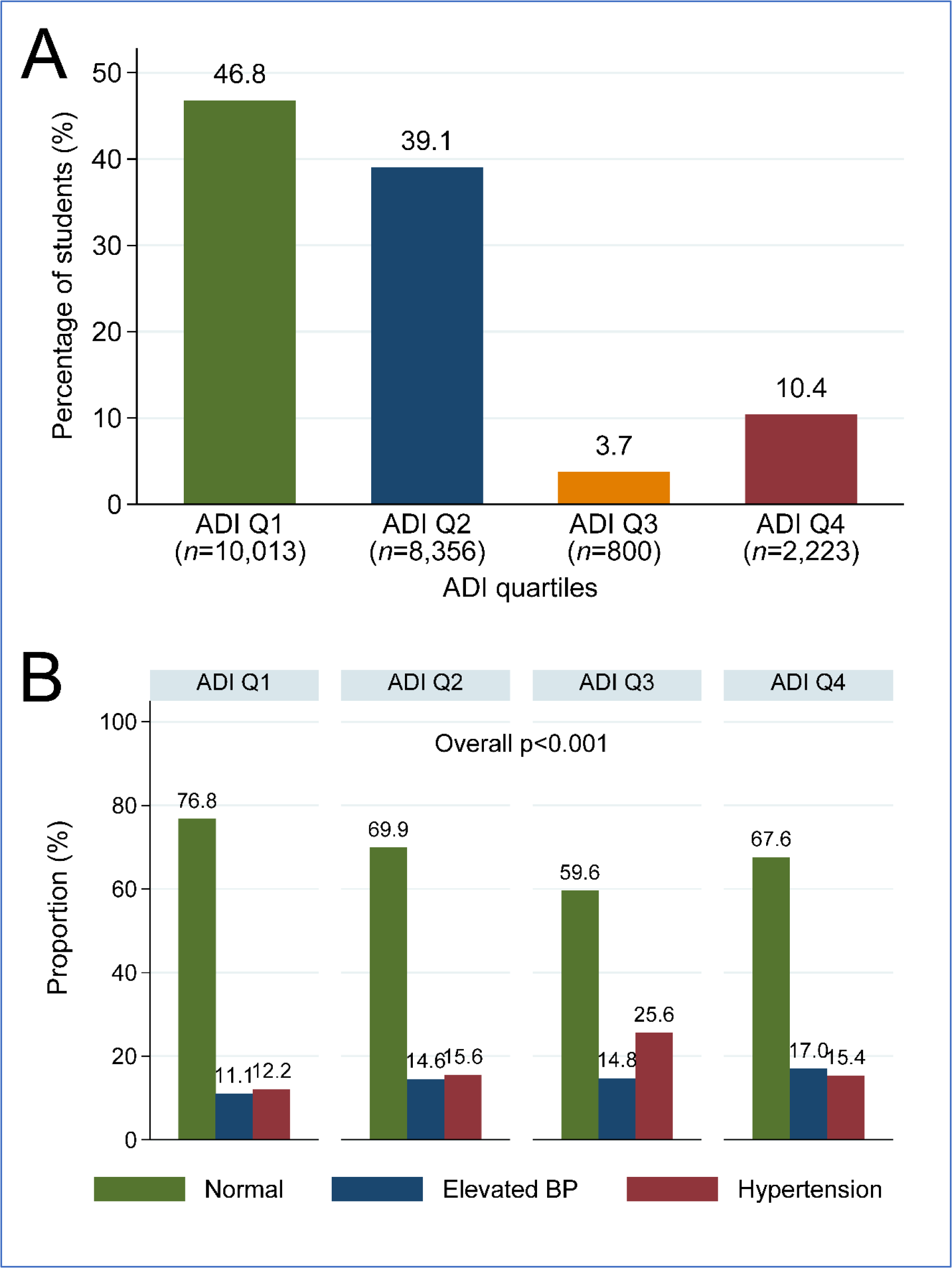
Distribution of students (A) and BP status (B) by schools’ weighted ADI quartiles

**Table 1:** Baseline characteristics by schools’ weighted Area deprivation Index (ADI)

|  | **Total**<br>**(N=21,392)** | **ADI Q1-2**<br>**(n=18,369)** | **ADI Q3-4**<br>**(n=3,023)** | **p-**<br>**value** |
| --- | --- | --- | --- | --- |
| Weighted ADI percentile, median (IQR) | 25.9 (12.7, 36.8) | 18.6 (11.4, 35.7) | 94.4 (73.5, 95.2) | <0.001 |
| Age (years), median (IQR) | 13.0 (12.0, 14.0) | 13.0 (12.0, 14.0) | 14.0 (13.0, 15.0) | <0.001 |
| Gender |  |  |  | 0.60 |
| Female | 11,006 (51.4) | 9,464 (51.5) | 1,542 (51.0) |  |
| Male | 10,386 (48.6) | 8,905 (48.5) | 1,481 (49.0) |  |
| Race/Ethnicity |  |  |  | <0.001 |
| White | 7,058 (33.2) | 6,870 (37.6) | 188 (6.2) |  |
| African American | 5,285 (24.8) | 3,614 (19.8) | 1,671 (55.5) |  |
| Hispanic | 7,179 (33.7) | 6,093 (33.4) | 1,086 (36.1) |  |
| Asian | 1,460 (6.9) | 1,424 (7.8) | 36 (1.2) |  |
| Other | 297 (1.4) | 267 (1.5) | 30 (1.0) |  |
| Weight z-score percentile, median (IQR) | 73.6 (45.7, 91.8) | 73.1 (45.4, 91.3) | 76.8 (47.9, 94.3) | <0.001 |
| BMI (kg/m<sup>2</sup>), median (IQR) | 21.3 (18.8, 25.0) | 21.1 (18.7, 24.6) | 22.8 (19.9, 27.7) | <0.001 |
| BMI z-score percentile, median (IQR) | 75.5 (48.1, 92.6) | 74.5 (46.9, 91.8) | 81.3 (55.0, 95.7) | <0.001 |
| BMI z-score percentile, categories |  |  |  | <0.001 |
| Normal Weight | 13,357 (62.4) | 11,690 (63.6) | 1,667 (55.1) |  |
| Overweight | 3,888 (18.2) | 3,354 (18.3) | 534 (17.7) |  |
| Obesity | 4,147 (19.4) | 3,325 (18.1) | 822 (27.2) |  |
| BP status (AAP) |  |  |  | <0.001 |
| Normal | 15,505 (72.5) | 13,525 (73.6) | 1,980 (65.5) |  |
| Elevated BP | 2,822 (13.2) | 2,326 (12.7) | 496 (16.4) |  |
| Hypertension | 3,065 (14.3) | 2,518 (13.7) | 547 (18.1) |  |
| Sustained Hypertension | 560 (2.6) | 422 (2.3) | 138 (4.6) | <0.001 |
| Language |  |  |  | <0.001 |
| English | 18,091 (84.6) | 16,818 (91.6) | 1,273 (42.1) |  |
| Non-English | 3,301 (15.4) | 1,551 (8.4) | 1,750 (57.9) |  |

**Table 2.** Characteristics associated with hypertension - Hypertension vs. Normal.

|  | Normal<br>(n=15,505) | Hypertension<br>(n=3,065) | Univariable |  |
| --- | --- | --- | --- | --- |
|  |  |  | OR (95% CI) | p-value |
| Age (years), median (IQR) | 13 (12, 14) | 13 (12, 14) | 1.05 (0.98, 1.11) | 0.15 |
| Gender |  |  |  |  |
| Female | 8,654 (55.8) | 1,309 (42.7) | (reference) |  |
| Male | 6,851 (44.2) | 1,756 (57.3) | 1.69 (1.42, 2.03) | <0.001 |
| Race/Ethnicity |  |  |  |  |
| White | 5,312 (34.5) | 912 (29.9) | (reference) |  |
| African American | 3,744 (24.3) | 748 (24.5) | 1.16 (0.92, 1.48) | 0.21 |
| Hispanic | 5,041 (32.7) | 1,182 (38.8) | 1.37 (1.09, 1.70) | 0.01 |
| Asian | 1,111 (7.2) | 172 (5.6) | 0.90 (0.75, 1.09) | 0.29 |
| Other | 211 (1.4) | 36 (1.2) | 0.99 (0.73, 1.36) | 0.97 |
| BMI (kg/m2), median (IQR) | 20.8 (18.5, 23.9) | 23.8 (20.0, 29.0) | 1.10 (1.09, 1.12) | <0.001 |
| BMI z-score percentile, median (IQR) | 71.6 (44.1, 90.0) | 89.0 (64.3, 97.6) | 1.02 (1.02, 1.02) | <0.001 |
| BMI z-score percentile, categories |  |  |  |  |
| Normal Weight | 10,462 (67.5) | 1,370 (44.7) | (reference) |  |
| Overweight | 2,784 (18.0) | 530 (17.3) | 1.45 (1.28, 1.66) | <0.001 |
| Obesity | 2,259 (14.6) | 1,165 (38.0) | 3.94 (3.51, 4.41) | <0.001 |
| Language |  |  |  |  |
| English | 13,253 (85.5) | 2,522 (82.3) | (reference) |  |
| Non-English | 2,252 (14.5) | 543 (17.7) | 1.27 (0.99, 1.62) | 0.06 |
| Number of fast-food store w. 0.5 mile, median (IQR) | 1.00 (0.00, 4.00) | 1.00 (0.00, 3.00) | 0.98 (0.96, 1.01) | 0.16 |
| Number of fast-food store w. 0.5-1 mile, median (IQR) | 1.00 (0.00, 4.00) | 1.00 (0.00, 3.00) | 0.98 (0.96, 1.01) | 0.16 |
| Weighted percent tree canopy, median (IQR) | 2.4 (1.3, 8.1) | 2.6 (1.3, 10.5) | 1.02 (1.00, 1.03) | 0.07 |
| Weighted ADI percentile, median (IQR) | 25.9 (12.3, 36.8) | 33.6 (16.5, 36.8) | 1.01 (1.00, 1.01) | 0.01 |
| Schools' weighted ADI |  |  |  |  |
| Q1 | 7,685 (49.6) | 1,218 (39.7) | (reference) |  |
| Q2 | 5,840 (37.7) | 1,300 (42.4) | 1.40 (1.11, 1.78) | 0.01 |
| Q3 | 477 (3.1) | 205 (6.7) | 2.71 (2.10, 3.50) | <0.001 |
| Q4 | 1,503 (9.7) | 342 (11.2) | 1.44 (1.17, 1.76) | <0.001 |
| Schools' weighted ADI |  |  |  |  |
| Q1-2 | 13,525 (87.2) | 2,518 (82.2) | (reference) |  |
| Q3-4 | 1,980 (12.8) | 547 (17.8) | 1.48 (1.10, 2.01) | 0.01 |
| Schools' weighted ADI |  |  |  |  |
| Q1 | 7,685 (49.6) | 1,218 (39.7) | (reference) |  |
| Q2-4 | 7,820 (50.4) | 1,847 (60.3) | 1.49 (1.20, 1.85) | <0.001 |

IQR, interquartile range; OR, odds ratio; CI, confidence interval; AUC, area under the receiver operating characteristic curve.

### Hypertension Prevalence based on Weighted School ADI

The largest proportion of students with normal BP was found among those attending schools in Q1. The prevalence of hypertension differed significantly across quartiles: 12.2% (1,218/10,013) in Q1, 15.6% (1,300/8,356) in Q2, 25.6% (205/800) in Q3 and 15.4% (342/2,223) in Q4 (p < 0.001) (Figure 1B).

The weighted ADI was significantly higher in the hypertension group, with a median of 33.6 (IQR 16.5, 36.8) compared to the group with normal BP, with a median of 25.9 (IQR 12.3, 36.8) (p=0.01). The highest prevalence of hypertension was observed in Q3 for all racial/ethnic groups: White (27.1%), Hispanic (25.6%), African American (21.2%), and Asian (22.2%). The highest rate of sustained hypertension at follow-up after two screens was observed in adolescents attending schools in Q3 and Q4, with 4.6% (136/3023), compared to 2.3% (422/18,369) in Q1 and Q2 (p < 0.001).

The prevalence of primary hypertension was highest among Hispanic adolescents (38.8%) followed by White (29.9%), African American (24.5%), Asian (5.6%), and other (1.2%). Compared to White adolescents, Hispanic students had a 37% increased risk of hypertension (OR 1.37; 95% CI, 1.09- 1.48; p=0.01) (Table 2). Except for Q3 where White students had the highest prevalence of hypertension. Hispanic students had the highest prevalence of hypertension in the remaining quartiles, followed by African American students, suggesting the genetic contribution to their hypertension risk persisted irrespective of socioeconomic factors (Figure 2A).

**Figure 2.**
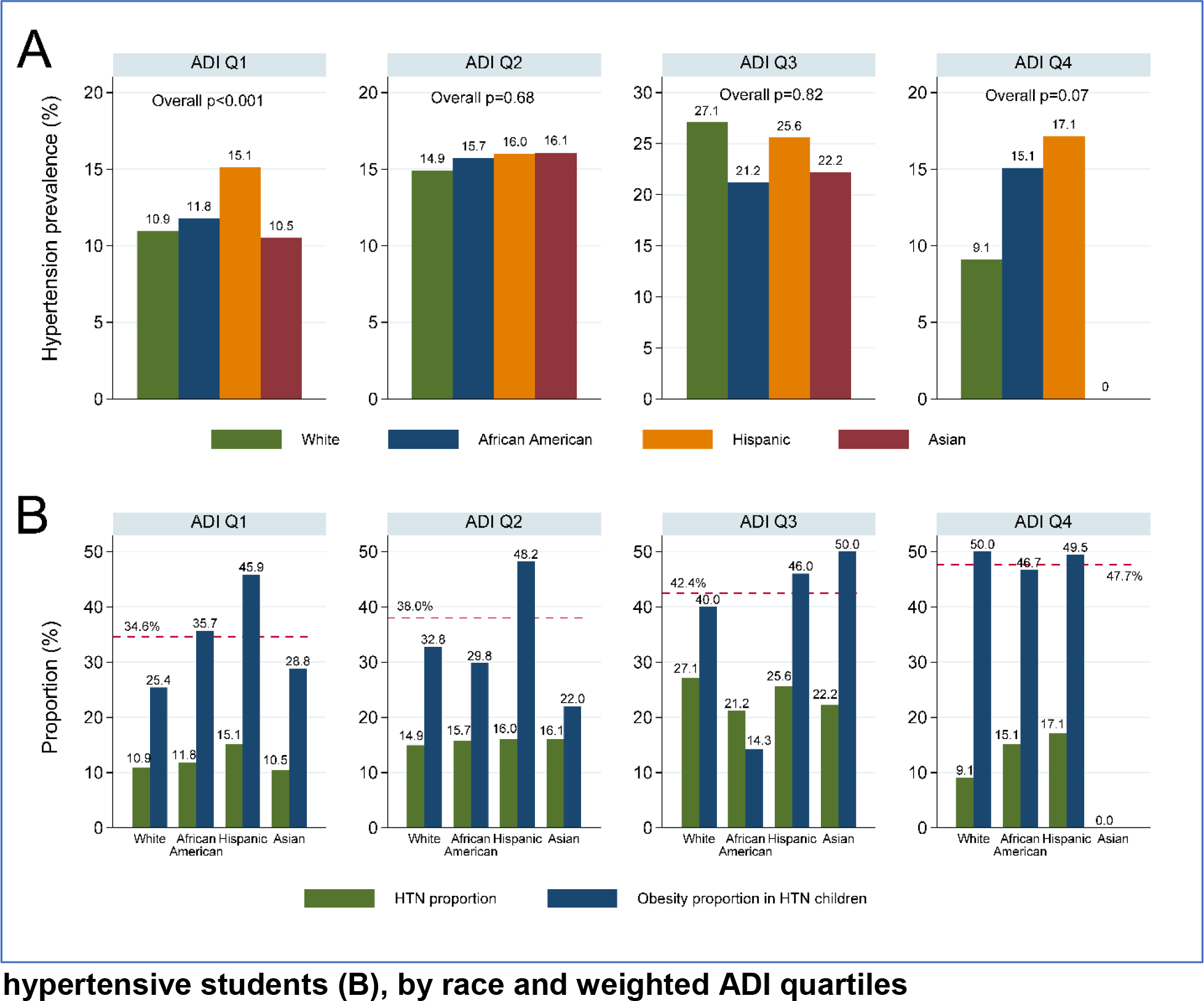
Distribution of hypertension (A) and hypertension with obesity among hypertensive students (B), by race and weighted ADI quartiles

There was a higher prevalence of obesity among the adolescents with hypertension compared to those with normal BP (38% vs. 14.6%; p< 0.001). Adolescents with hypertension were 3.88 times more likely to be obese compared to those with normal BP (Table 2). The prevalence of obesity increased with social deprivation with the lowest rates seen in Q1 and the highest rates in Q4. Hispanic adolescents had the highest obesity rates in each quartile, significantly higher than the quartile average rate. These high obesity rates might have influenced the rates of hypertension seen in Hispanic children across different quartiles, especially in Q3. The influence of obesity on hypertension risk for White adolescents was seen in Q3. (Figure 2B).

In multivariable logistic regression analysis, being male, multiracial, overweight, obese, and attending school in Q3 was significantly associated with sustained hypertension (Table 3). Compared to attending schools located in affluent neighborhoods (Q1), adolescents attending a school in Q2-Q4 had more than a 30% increase in the odds of having sustained hypertension [OR 1.33 (95% CI, 1.08, 1.65); p=0.01] (Figure 3).

**Figure 3:**
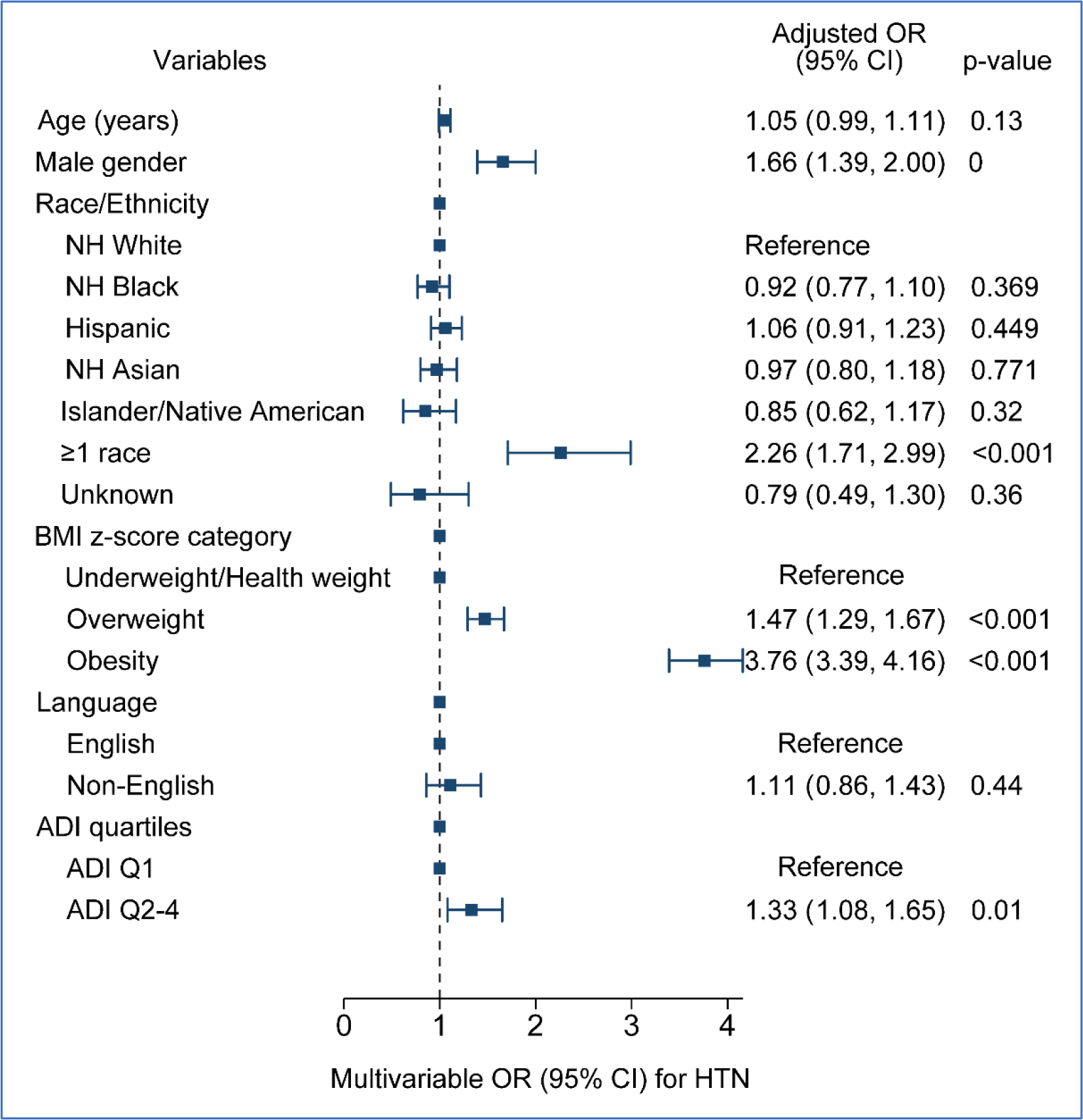
Forest plot - Factors associated with hypertension, multivariable logistic regression, ADI Q2-4 vs. Q1

**Table 3.** Characteristics associated with hypertension, multivariable logistic regression - Hypertension vs. Normal.

regression - Hypertension vs. Normal
|  | 3.1. ADI quartiles |  | 3.2. ADI Q3-4 vs. Q1-2 |  |
| --- | --- | --- | --- | --- |
|  | Multivariable |  | Multivariable |  |
|  | OR (95% CI) | p-value | OR (95% CI) | p-value |
| Schools' weighted ADI |  |  |  |  |
| Q1 | (reference) |  | -- | -- |
| Q2 | 1.31 (1.03, 1.66) | 0.03 | -- | -- |
| Q3 | 2.24 (1.49, 3.36) | <0.001 | -- | -- |
| Q4 | 1.12 (0.90, 1.38) | 0.31 | -- | -- |
| Schools' weighted ADI |  |  |  |  |
| Q1-2 | -- | -- | (reference) |  |
| Q3-4 | -- | -- | 1.30 (0.87, 1.95) | 0.21 |
| Age (years) | 1.05 (1.00, 1.11) | 0.06 | 1.05 (1.00, 1.10) | 0.03 |
| Male gender | 1.66 (1.38, 1.99) | <0.001 | 1.66 (1.39, 1.98) | <0.001 |
| Race/Ethnicity |  |  |  |  |
| White | (reference) |  | (reference) |  |
| African American | 0.98 (0.81, 1.19) | 0.85 | 0.92 (0.69, 1.22) | 0.56 |
| Hispanic | 1.04 (0.89, 1.21) | 0.62 | 1.09 (0.90, 1.32) | 0.39 |
| Asian | 0.98 (0.81, 1.20) | 0.87 | 0.94 (0.76, 1.15) | 0.54 |
| Other | 0.84 (0.61, 1.15) | 0.28 | 0.85 (0.61, 1.20) | 0.37 |
| BMI z-score percentile, categories |  |  |  |  |
| Normal Weight | (reference) |  | (reference) |  |
| Overweight | 1.47 (1.29, 1.67) | <0.001 | 1.47 (1.29, 1.68) | <0.001 |
| Obesity | 3.78 (3.42, 4.18) | <0.001 | 3.81 (3.45, 4.21) | <0.001 |
| Language |  |  |  |  |
| English | (reference) |  | (reference) |  |
| Non-English | 1.08 (0.84, 1.38) | 0.57 | 0.97 (0.75, 1.26) | 0.83 |

OR, odds ratio; CI, confidence interval

## Discussion

This study investigated the relationship between neighborhood deprivation and hypertension in a large, diverse sample of adolescents in Houston, Texas. Our findings reveal a concerning association between higher neighborhood deprivation and increased prevalence of hypertension, independent of race/ethnicity highlights the influence of social and environmental factors in disadvantaged communities on hypertension prevalence among adolescents. This association was seen across all ethnicities, except Asian students. However, attending schools in affluent neighborhoods appeared to lower risk of hypertension, possibly due to healthier food options, green spaces, and more opportunities for physical activity than students who reside in deprived neighborhoods.

Racial and ethnic minority adolescents, particularly Hispanic and African American adolescents, are disproportionately affected by primary hypertension. Schools in socially disadvantaged neighborhoods had a disproportionately higher attendance rate of Hispanic and African American adolescents, contributing to the elevated hypertension rates in these areas. These disparities are likely rooted in discriminatory housing practices like redlining, which resulted in racial/ethnic minorities living in socially disadvantaged neighborhoods with limited access to resources. ^36^ These environmental factors contribute to higher rates of chronic diseases and poorer health outcomes in these populations.^37^ Our findings are consistent with a recent cross-sectional analysis in North Carolina by Mohottige et al., which identified an increased burden of structural racism indicators associated with higher neighborhood prevalence of hypertension, including lower percentages of White individuals in the neighborhood, high poverty rates, and higher ADI. ^38^

The association between neighborhood deprivation and hypertension risk appears to be partially mediated by obesity, particularly in White and Hispanic adolescents. The influence of social deprivation was seen in White obese adolescents who had an increased risk for hypertension when they attended schools in disadvantaged neighborhoods. Adolescents attending schools in impoverished neighborhoods have higher rates of obesity likely related to poor dietary behaviors and limited physical activity opportunities. There is growing literature showing the effects of neighborhoods on dietary behavior and physical activity. Several neighborhood characteristics including unfavorable food environments, poor walkability, and high crime rates make active, healthy living in disadvantaged neighborhoods difficult.

Although Hispanic adolescents in socially deprived neighborhoods had a significantly high risk for hypertension, they were also noted to have higher hypertension rates compared to other racial/ethnic groups when attending schools in affluent neighborhoods suggesting the influence of genetic and cultural factors. However, the contribution of genetics to ethnic differences in hypertension prevalence remains highly controversial.^39^ Social and cultural factors rather than genetics are thought to explain differences in hypertension prevalence between Hispanics and other ethnic groups in the United States emphasizing the importance of social determinants of hypertension risk in the Hispanic population.

The well-documented tracking of elevated BP into adulthood^3^ and the increased risk for chronic kidney disease associated with hypertension, ^40^ highlight the importance of identifying modifiable risk factors for primary hypertension in adolescents.

Socioeconomic factors play a significant role in hypertension prevalence, drawing attention to the social drivers of hypertension risk. Individuals who live in socially disadvantaged neighborhoods often experience higher rates of food insecurity, which is linked to consumption of salty, energy-dense foods, increasing the risk of obesity and hypertension. Addressing food insecurity through policy changes and clinical interventions could potentially mitigate hypertension risk.

Our study has several limitations. Its cross-sectional nature limits our ability to establish causality or discern temporal relationships between potential risk factors and outcomes. Additionally, using school ADI as a proxy for students’ neighborhoods may oversimplify socioeconomic conditions, and other individual-levels factors may contribute to hypertension risk. Moreover, the inclusion of only two private schools which are not affected by residential zoning regulation may limit the generalizability of our findings. Lastly, the use of BMI for weight classifications may have overestimated body fat in certain populations such as athletes and students with a muscular build.

Despite these limitations, our study benefits from comprehensive data capturing a diverse socioeconomic and racial/ethnic sample representative of the Houston population. This school-based study enabled participation from children with limited healthcare access, reducing selection bias often seen in clinic-based studies.

Furthermore, the study utilizes a large and diverse sample, allowing for robust analysis and making statistically compelling comparisons between different socioeconomic groups.

In conclusion, our findings highlight the critical role of social determinants of health in shaping adolescents’ cardiovascular health. It demonstrates that attending school in neighborhoods with lower socioeconomic status contributes to racial disparities in hypertension risk. Schools in disadvantaged neighborhoods could implement targeted programs promoting healthy lifestyles and mitigating environmental risk factors. There is a need for targeted public health initiatives addressing access to healthy foods, physical activity opportunities, and quality healthcare in underserved communities. Additionally, early identification and management of modifiable risk factors like obesity in high-deprivation areas are crucial. Longitudinal studies are needed to explore the causal pathways linking neighborhood deprivation and hypertension risk. There is also a need to investigate specific social and environmental factors further within disadvantaged communities that contribute to hypertension. Additionally, the findings underscore the importance of advocating for policy changes to ensure equitable school environments and reduce hypertension disparities. By addressing social determinants of health alongside individual risk factors, we can create a more comprehensive approach to preventing hypertension in adolescents, particularly those residing in disadvantaged communities.

### Perspectives

Adolescents who attend schools in impoverished neighborhoods have a greater risk for hypertension highlighting the influence of neighborhood and built environment on hypertension risk. Constrained by housing policies, there was a disproportionate representation of Hispanic and African American adolescents in the high-poverty school neighborhoods. Among racial/ethnic groups, Hispanic students had the highest prevalence of sustained hypertension across ADI quartiles followed by African American students. The effect of school attendance in impoverished neighborhoods was also observed in White obese students. Our findings have significant public health implications and shed light on the racial and socioeconomic disparities that exist in hypertension prevalence. Our study emphasizes the importance of implementing targeted interventions to address these issues including dietary intervention and physical activities within the school environment.

## NOVELTY AND RELEVANCE

### What Is New?

Using a comprehensive dataset capturing a diverse socioeconomic and racial/ethnic student population we demonstrated the influence of the school environment on hypertension risk utilizing a validated measure of socioeconomic status to categorize neighborhood deprivation.

### What Is Relevant?

Adolescents attending school in high-poverty school environments have a higher risk of hypertension

### Clinical/Pathophysiological Implications?

Our findings highlight the critical impact of the school environment on hypertension risk. We provide evidence to support public health initiatives including targeted dietary and physical activity interventions to reduce the racial and socioeconomic disparities in hypertension risk.

## Data Availability

Data is available from University of Texas Health Science Center at Houston on request and by IRB approval

## Acknowledgments Sources of Funding

This work was funded, in part, by federal funds from the USDA/ARS under Cooperative Agreement no. 58-3092-0-001 (J.M.D.). The contents of this publication do not necessarily reflect the views or policies of the USDA nor does the mention of trade names, commercial products, or organizations imply endorsement from the U.S. government.

## Disclosures

None

## Notes

### Competing Interest Statement

The authors have declared no competing interest.

### Clinical Trial

N/A

### Author Declarations

Institutional Review Board at the University of Texas Health Science Center at Houston

